# Carsickness Therapy Based on Brain–Computer Interface Enhanced Mindfulness Meditation Training

**DOI:** 10.64898/2026.04.01.26349963

**Authors:** Jiawei Zhu, Zhenfu Wen, Yonghao Cao, Qiyun Huang, Yuanqing Li

## Abstract

Carsickness impairs comfort and affects a large proportion of the population. However, interventions that provide a therapeutic solution to carsickness have yet to be established. Here we introduce a wearable mindfulness meditation brain–computer interface (MM-BCI) system as a closed-loop training therapy for carsickness. The system records electroencephalographic activity, decodes meditative state in real time and delivers audiovisual neurofeedback to scaffold meditation practice. In a 10-week randomized controlled trial, 60 individuals susceptible to carsickness were assigned to practice mindfulness meditation with either real-time MM-BCI neurofeedback or sham feedback, both during real-world car riding and at home. Critically, pre-intervention, post-intervention, and one-month follow-up assessments of carsickness severity were conducted during regular car riding without any task or feedback system. Relative to the sham group, the MM-BCI group showed significantly reduced carsickness severity at post-intervention and follow-up. At baseline, carsickness-susceptible participants exhibited a reduced aperiodic exponent in occipito-parietal cortex relative to non-susceptible controls, identifying a candidate neural signature of carsickness susceptibility. MM-BCI training increased this exponent toward non-susceptible levels, and the magnitude of this neural normalization was associated with the degree of symptom improvement. This study provides the first demonstration that BCI-enhanced mindfulness meditation can induce promising treatment effect on carsickness, offering a transformative non-pharmacological approach to enhance passenger well-being in everyday transit.

## 1 Introduction

Motion sickness, specifically carsickness, is defined as a condition of feeling unwell that can occur when traveling in a car [1]. This phenomenon is most commonly explained by the sensory conflict theory, which posits that the discomfort arises from a mismatch between visual, vestibular, and proprioceptive signals regarding self-motion [2]. A large-scale international survey reveals that carsickness remains a prevalent issue, affecting approximately 46% of car passengers within a five-year period, and this lifetime incidence increases to about 59% when childhood experiences are included [3]. Therefore, it is imperative to establish effective intervention methods for carsickness.

Pharmacological treatments for carsickness can be effective but often cause sedation and other side effects, limiting their acceptability for frequent use [4]. Non-pharmacological strategies, such as providing predictive motion cues [5], [6], shifting attention away from bodily sensations [7], [8] or using intelligent vehicle control algorithms [9], [10], offer more acceptable alternatives. However, their practical efficacy often lacks rigorous validation in large-scale, real-world settings, and widespread implementation remains limited [2], [11], [12]. Moreover, most existing approaches are designed to relieve symptoms only during a given journey and therefore must be applied repeatedly for each ride. In our previous work, we developed a wearable mindfulness meditation brain–computer interface (MM-BCI) system, and demonstrated its effectiveness in producing immediate symptom relief in real-world car and maritime settings [13], [14]. Despite these advancements, a critical question remains: can BCI-based training move beyond temporary, ride-specific relief and produce therapeutic effect on carsickness? At present, an intervention that provides therapeutic effect on carsickness beyond the immediate period of use has yet to be established.

Mindfulness meditation comprises a family of cognitive and emotional training practices that cultivate attention to and acceptance of present-moment experiences [15], [16]. Sustained mindfulness meditation practice has been shown to improve attentional capacities, particularly in the domain of sustained attention and executive control [17], [18]. These enhanced capacities may support more efficient processing of the sensory conflicts [19], which is thought to underlie carsickness [2]. However, traditional mindfulness meditation can be difficult for novices to initiate and sustain. Novices often receive little objective feedback on whether they are practicing effectively, and the training can be monotonous and lacks tailored instruction, contributing to high attrition rates due to discouragement [20], [21]. Brain–computer interface (BCI) has the potential to enhance mindfulness meditation by providing a direct link between internal neural state and external feedback [22]. Building on this idea, we recently developed a MM-BCI system to alleviate carsickness and seasickness [13], [14]. The system operates via a wearable headband that acquires prefrontal electroencephalography (EEG) signals while the user engages in mindfulness meditation practice, such as focusing on breathing. These signals are analyzed in real-time to estimate user’s meditative state, and are then translated into dynamic audiovisual feedback. This closed-loop design enables users to sustainably shift their attention from the changing vehicular environment toward mindfulness meditation practices, which reduces the sensory mismatch and thereby alleviating symptoms. In contrast, traditional mindfulness meditation without such real-time feedback fails to achieve significant symptom relief.

Building on our previous studies demonstrating immediate symptom relief with MM-BCI, the present study tested whether this closed-loop approach can move beyond acute aid to produce treatment effect on carsickness. We conducted a 10-week randomized controlled trial using a real-world regular car riding without any task or feedback system at all assessments (pre-intervention, post-intervention, and one-month follow-up; Figure 1). The results demonstrated that MM-BCI training produced significant and sustained reductions in carsickness severity among susceptible individuals. Furthermore, EEG analysis identified the occipito-parietal aperiodic exponent, a measure derived from the slope of the power spectrum [23], as a candidate neural signature of carsickness susceptibility. Critically, MM-BCI training normalized this neural signature toward levels observed in non-susceptible individuals. Together, these findings demonstrate that BCI-enhanced mindfulness meditation can produce treatment effect on carsickness while reshaping neural dynamics associated with carsickness susceptibility.

**Figure 1.**
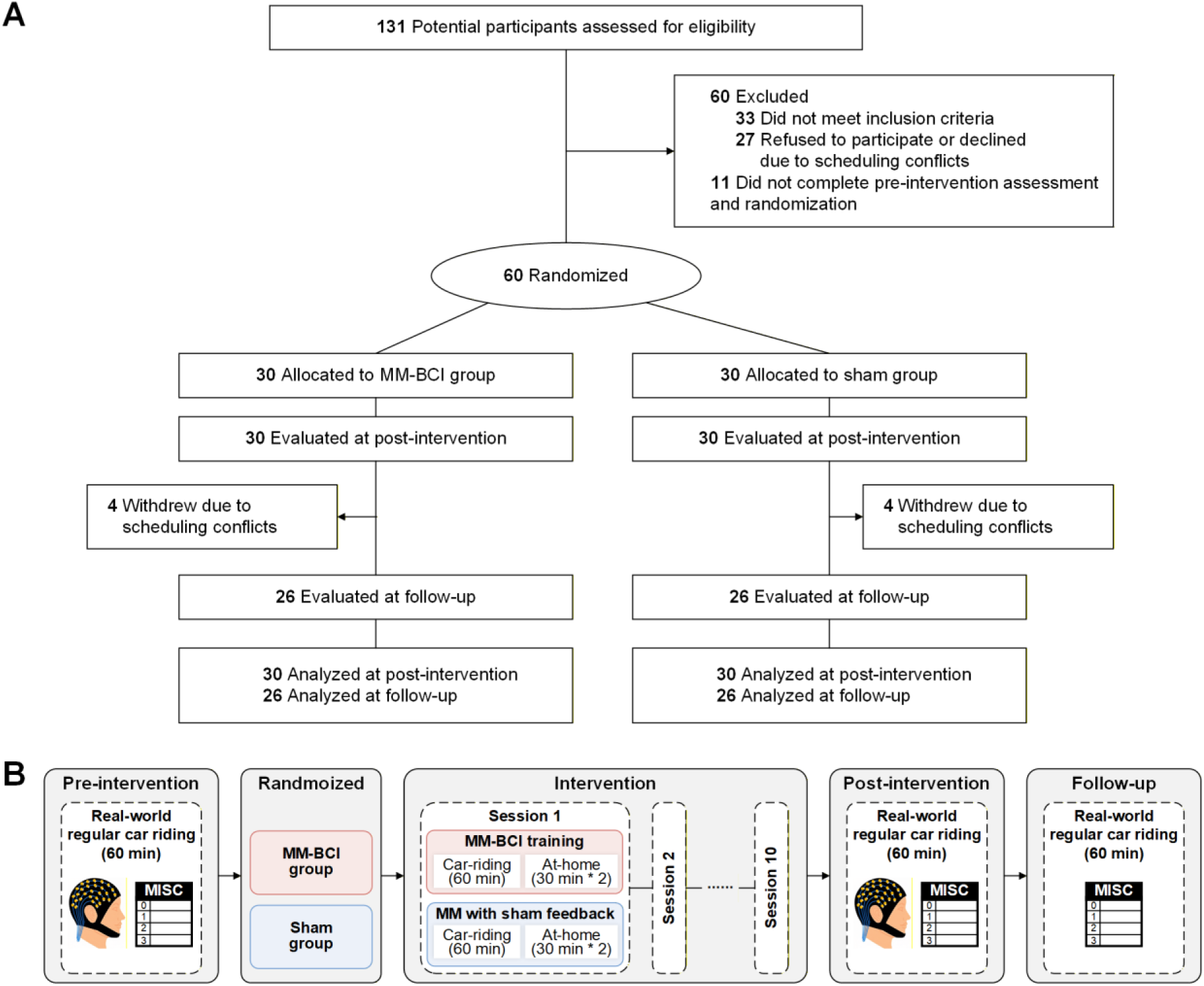
Participant disposition and study design.

## 2 Results

Sixty individuals susceptible to carsickness were randomly assigned to practice mindfulness meditation with either real-time MM-BCI neurofeedback (MM-BCI group, N=30) or sham feedback (sham group, N=30) during ten weekly one-hour real-world car rides, supplemented by at-home practice. A separate cohort of individuals non-susceptible to carsickness (non-carsickness group, N=31) was recruited to provide a baseline for neural comparisons. Primary outcomes included subjective carsickness severity (Misery Scale, MISC [24]) assessed during one-hour regular car riding without any task or feedback system at pre-intervention, post-intervention, and one-month follow-up. Electrophysiological measures, specifically the aperiodic exponent derived from EEG recordings (a 64-channel EEG cap), were obtained during both car-riding and resting states at pre- and post-intervention.

### 2.1 MM-BCI training yields significant treatment effect on carsickness

Following the post-intervention assessment, participants rated the perceived effectiveness of the 10-week training in treating carsickness on a 7-point Likert scale (1 = strongly agree to 7 = strongly disagree). A larger proportion of participants in the MM-BCI group (96.67%, 29/30; Figure 2A) reported a positive view compared to the sham group (76.67%, 23/30). An independent-samples t-test on the Likert scale scores confirmed that the MM-BCI group rated the training as significantly more effective than the sham group (MM-BCI: 2.100±0.121, sham: 2.700±0.174; t(51.712) = 2.834, P = 0.007, Cohen’d = 0.732). This result indicates that participants who received MM-BCI training held a more favorable subjective view of its benefit than those received sham feedback. Supporting this positive perception, most participants in the MM-BCI group reported that they could tolerate car riding with only mild and manageable symptoms even without using the system.

**Figure 2.**
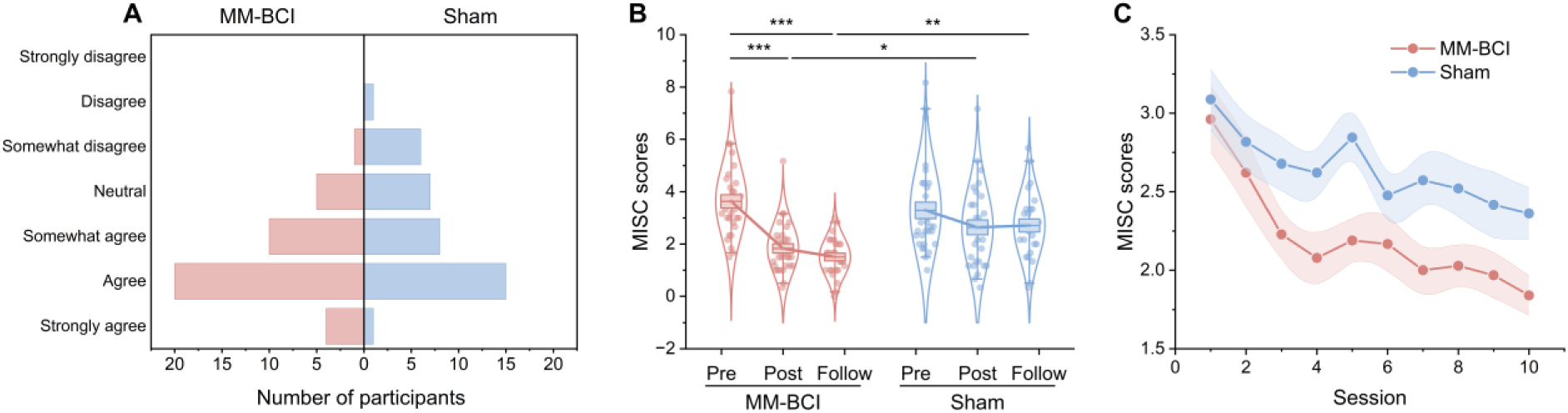
Behavioral outcomes across assessments and during the intervention period. A, Evaluation results for the effectiveness of the 10-week training for addressing carsickness. B, The averaged MISC scores of MM-BCI and sham groups in different assessments. C, Weekly MISC scores of both groups recorded during the 10-week car-riding meditation training sessions. The center line represents the mean, the box and shadow indicate the standard error of the mean (s.e.m), and the whiskers extend to the 5th and 95th percentiles. *P < 0.05, **P < 0.01, ***P < 0.001.

To assess the longitudinal efficacy of the intervention on carsickness severity, subjective symptom levels were assessed using the MISC, with lower scores indicating milder symptoms. A 2 (group: MM-BCI, sham) × 3 (time: pre, post, follow-up) repeated-measures ANOVA revealed a significant group × time interaction (F(2, 166) = 5.174, P = 0.007, ηp² = 0.059; Figure 2B). Simple effects analyses with Benjamini-Hochberg correction showed no significant group difference at pre-intervention (t(58) = 0.847, P = 0.400, Cohen’d = 0.219), indicating comparable baseline symptom severity. In contrast, the MM-BCI group reported significantly lower MISC scores than the sham group at both post-intervention (t(48.846) = - 2.362, P = 0.032, Cohen’d = -0.610) and at the one-month follow-up (t(50) = -4.165, P < 0.001, Cohen’d = -1.155). Within the MM-BCI group, a significant main effect of time was found (F(2, 83) = 31.844, P < 0.001, ηp² = 0.434). Post-hoc tests confirmed that MISC scores at both post-intervention (paired t(29) = -8.053, P < 0.001, Cohen’d = -1.470) and follow-up (paired t(25) = -8.840, P < 0.001, Cohen’d = -1.734) were significantly lower than at pre-intervention, with no significant difference between post-intervention and follow-up (paired t(25) = 1.365, P = 0.184), indicating that symptom reduction was maintained over time. In contrast, the sham group showed no significant main effect of time (F(2, 83) = 1.579, P = 0.212), suggesting no reliable change in carsickness severity across assessments.

To further characterize the trajectory of symptom change during the intervention, we analyzed weekly MISC scores assessed during the 10 car-riding training sessions. A two-way (group × session) repeated-measures ANOVA revealed a significant main effect of group (F(1, 580) = 16.402, P < 0.001, ηp² = 0.028), with the MM-BCI group consistently reporting lower symptom severity than the sham group across sessions (Figure 2C). A significant main effect of session was also observed (F(9, 580) = 2.627, P = 0.006, ηp² = 0.039). Post-hoc comparisons with Benjamini-Hochberg correction revealed that symptom severity was significantly lower than in session 1 from session 3 onward (all P< 0.01), reflecting a gradual reduction in symptoms over the course of training. The group × session interaction was not significant (F(9, 580) = 0.252, P = 0.986), indicating a stable advantage for the MM-BCI group across training sessions.

### 2.2 Occipito-parietal aperiodic exponent serves as a potential neural signature for carsickness susceptibility

Why does MM-BCI training provide a sustained behavioral benefit, and are there neurophysiological modifications underlying this effect? To begin addressing this question, we first asked whether carsickness-susceptible individuals share a common neurophysiological profile at baseline. Specifically, we examined baseline EEG aperiodic exponents in carsickness-susceptible and non-susceptible participants.

A cluster-based permutation test (one-way ANOVA model, electrode-wise threshold P = 0.01, F = 4.85) on the car-riding state revealed a significant occipito-parietal cluster comprising five electrodes (O1, Oz, PO3, PO5, PO7; F_cluster_(2, 88) = 7.127, P_cluster_ = 0.016, ηp^2^ = 0.161; ^F^_igure_ ^3^A). Post-hoc independent-samples t-tests with Benjamini-Hochberg correction on the mean component within this cluster showed no significant difference between the two carsickness groups (MM-BCI vs. sham: t(58) = 1.721, P = 0.091; Figure 3B). In contrast, both the MM-BCI group (t(59) = -2.284, P = 0.039, Cohen’d = -0.585) and the sham group (t(59)= -4.340, P < 0.001, Cohen’d = -1.111) exhibited significantly lower aperiodic exponent than the non-carsickness group, indicating a flatter aperiodic slope in occipito-parietal cortex among carsickness-susceptible individuals.

**Figure 3.**
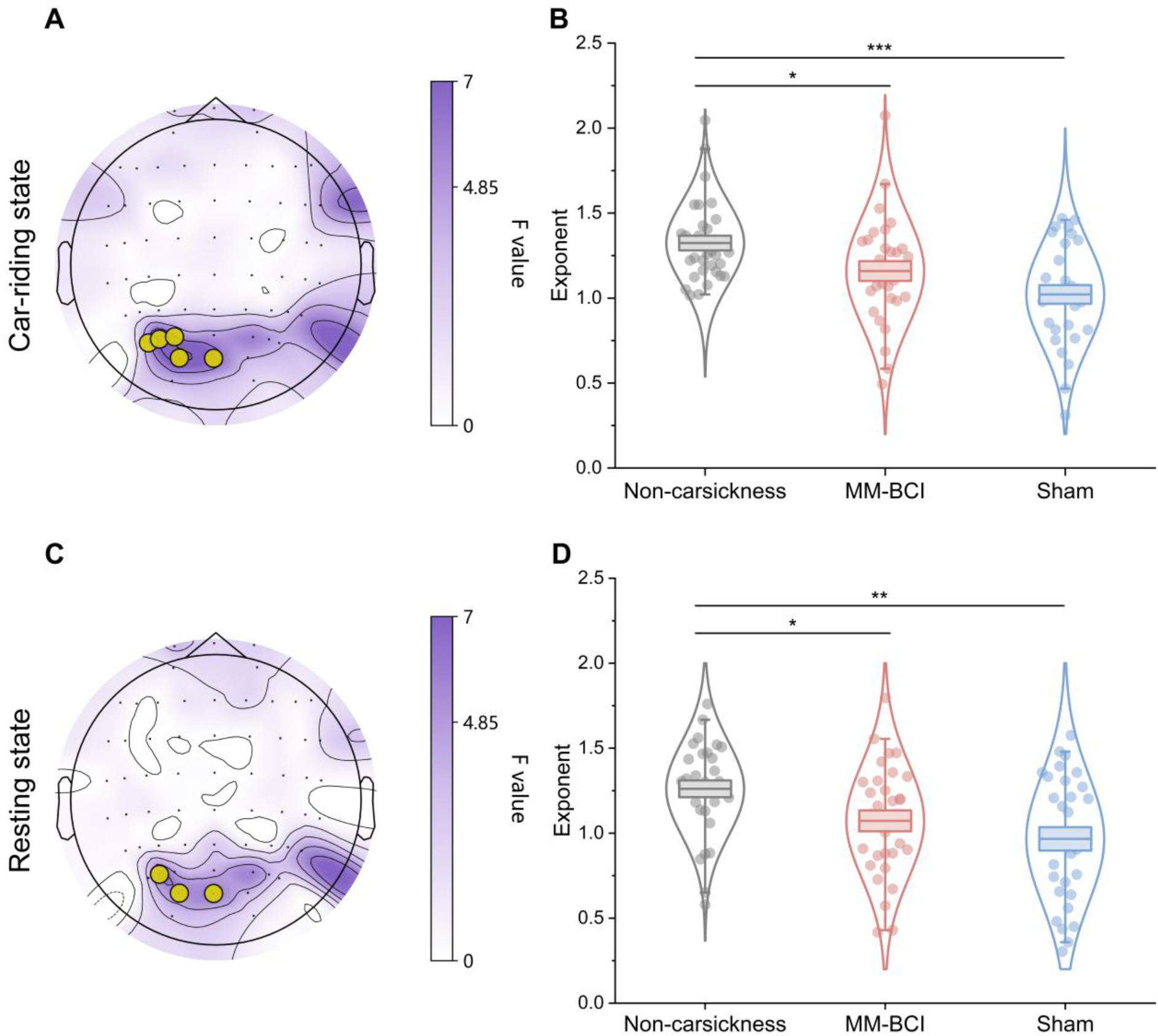
Baseline (pre-intervention) differences in aperiodic exponent associated with carsickness susceptibility. A, Topographic map of the F-statistic from a one-way ANOVA comparing the aperiodic exponent across the non-carsickness, MM-BCI, and sham groups during the car-riding state. Electrodes forming a significant cluster (O1, Oz, PO3, PO5, PO7) after cluster-based permutation correction (P_cluster_ = 0.016, ηp^2^ = 0.161) are outlined in yellow circle. B, The mean aperiodic exponent averaged across the significant electrode cluster identified in (A) for individuals in each group during the car-riding state. C, D, Corresponding topographic map of the F-statistic (C) and group-level boxplots (D) for the aperiodic exponent during the resting state. The center line represents the mean, the box indicates s.e.m, and the whiskers extend to the 5th and 95th percentiles. *P < 0.05, **P < 0.01, ***P < 0.001.

Applying the same analysis to the resting-state data revealed a spatially similar significant occipito-parietal cluster (O1, Oz, PO5; F_cluster_(2, 88) = 5.736, P_cluster_= 0.040, ηp^2^ = 0.123; Figure ^3^C). Similarly, no significant difference was observed between the two carsickness groups (MM-BCI vs. sham: t(58) = 1.150, P = 0.255; Figure 3D), while both carsickness groups showed significantly reduced aperiodic exponents relative to the non-carsickness group (MM-BCI vs. non-carsickness: t(59)= -2.401, P = 0.029, Cohen’d = -0.615; Sham vs. non-carsickness: t(59)= -3.499, P = 0.003, Cohen’d = -0.896). These results indicate that reduced occipito-parietal aperiodic exponent is a stable baseline neurophysiological signature of carsickness-susceptible individuals, present both at rest and during car riding.

### 2.3 MM-BCI normalizes occipito-parietal aperiodic exponent in association with symptom reduction

Having identified that reduced occipito-parietal aperiodic exponent characterizes carsickness-susceptible individuals at baseline, we next examined whether MM-BCI training modifies this neural signature and whether such changes relate to symptom improvement.

During the car-riding state, a one-way ANOVA on post-intervention aperiodic exponents revealed a significant group effect (F(2, 88) = 6.304, P = 0.003, ηp² = 0.125; Figure 4A). Post-hoc independent-samples t-tests with Benjamini-Hochberg correction showed that the MM-BCI group had a significantly higher (more normalized) aperiodic exponent than the sham group (t(58) = 2.812, P = 0.010, Cohen’d = 0.726), and was statistically indistinguishable from the non-carsickness group (t(59) = 0.129, P = 0.898). In contrast, aperiodic exponent of the sham group remained significantly reduced relative to the non-carsickness group (t(59) = - 2.900, P = 0.010, Cohen’d = -0.743). Within-group comparisons from pre- to post-intervention confirmed this pattern. Paired-samples t-tests showed a significant increase in aperiodic exponent in the MM-BCI group (t(29) = 3.254, P = 0.003, Cohen’s d = 0.594; Figure 4B), whereas the sham group showed no significant change (t(29) = 0.823, P = 0.417).

**Figure 4.**
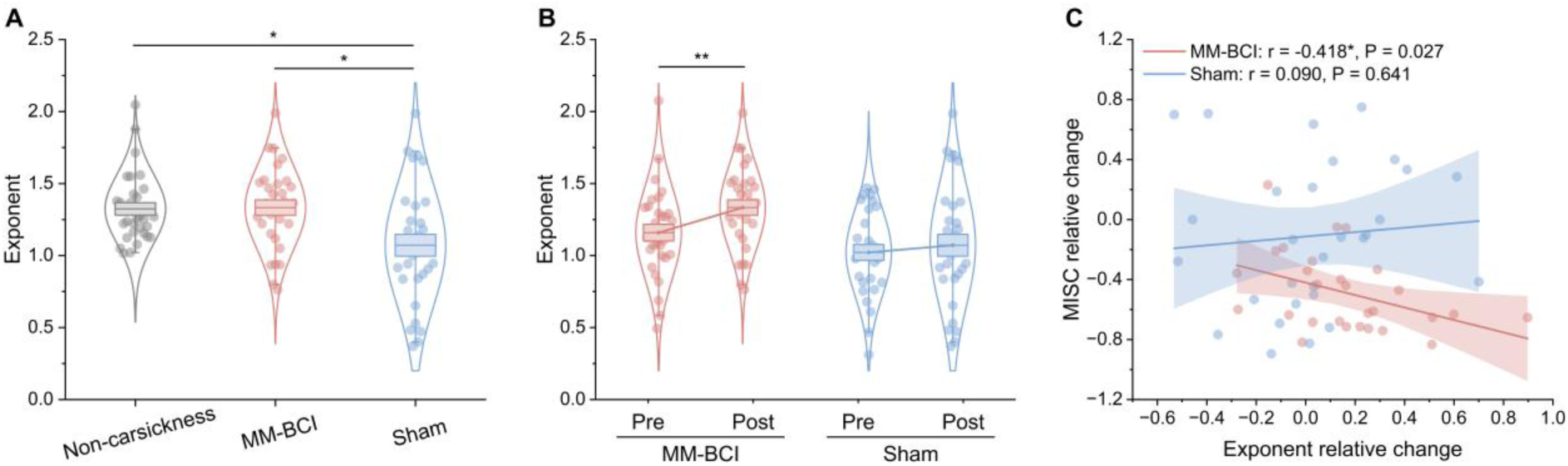
Post-intervention results of the occipito-parietal aperiodic exponent during the car-riding state. A, Mean aperiodic exponent (averaged across significant electrode cluster O1, Oz, PO3, PO5, PO7) for the MM-BCI and sham groups at post-intervention assessment against the baseline of non-carsickness group. B, Mean aperiodic exponent for the MM-BCI and sham groups at pre- and post-intervention assessment. C, Correlation between the relative change ((post-intervention – pre-intervention)/pre-intervention) in the aperiodic exponent and the relative change in MISC score for the car-riding state. The center line represents the mean, the box indicates s.e.m, and the whiskers extend to the 5th and 95th percentiles. *P<0.05, **P < 0.01.

To examine the relationship between neural plasticity and symptom reduction, we correlated relative changes ((post − pre)/pre) in aperiodic exponent with relative changes in MISC scores over the intervention period. Three participants (2 in the MM-BCI group, 1 in the sham group) were excluded from this analysis because their relative change values exceeded ±2.5 s.d. from the group mean. Pearson correlation analysis for the car-riding state (Figure 4C) revealed a significant negative correlation in the MM-BCI group (r= -0.418, P = 0.027): larger increases in occipito-parietal aperiodic exponent were associated with greater reductions in carsickness severity. No such relationship was observed in the sham group (r= 0.090, P = 0.641).

## 3 Discussion

This randomized controlled trial demonstrates that mindfulness meditation brain–computer interface (MM-BCI) training is an effective carsickness therapy, producing significant treatment effect on carsickness in real-world settings. After 10 weeks of training, participants in the MM-BCI group showed a robust reduction in carsickness severity that was maintained at the one-month follow-up, an effect not observed in the sham group. At the neural level, we identified the aperiodic exponent in occipito-parietal cortex as a potential neurophysiological signature of carsickness susceptibility and showed that MM-BCI training significantly normalized this aberrant neural activity. To our knowledge, the present study is the first to demonstrate that MM-BCI training can produce promising therapeutic effect on carsickness under real-world car rides [2].

Compared to pre-intervention, the MM-BCI group exhibited a significant reduction in carsickness symptoms at both post-intervention and follow-up, whereas the sham group showed no significant changes across assessments (Figure 2B). These results suggest that the improvement in the MM-BCI group was attributable to the neurofeedback itself rather than nonspecific factors such as placebo effects. This efficacy can be explained by the distinct training processes. During the training sessions, participants in the MM-BCI group could continuously redirect their attention from carsickness discomfort to mindfulness meditation based on real-time, valid neurofeedback, leading to immediate symptom relief (Figure 2C) [13]. In contrast, the sham group, receiving feedback unrelated to their actual state, showed poorer cognitive adjustment (reflected in lower meditation scores and a less favorable frontal EEG profile, i.e., a higher theta/alpha ratio [25], [26]; Figure S1). Specifically, the MM-BCI system provided real-time, objective audiovisual feedback on meditative state, which not only helped users promptly detect and correct mind-wandering but also made mindfulness practice more accessible, engaging, and sustainable. These advantages were not afforded by either traditional meditation or sham feedback, as both lacked such contingent guidance that enhance motivation and learning efficiency [20], [21]. Over repeated training, the MM-BCI group internalized the ability to disengage attention from discomfort, resulting in a significant treatment effect that persisted even without the device. The sham group, lacking correct feedback guidance, failed to acquire this skill and showed no such therapeutic improvement.

The occipito-parietal aperiodic exponent was identified as a neural signature of carsickness susceptibility, being significantly lower in both MM-BCI and sham groups compared to the non-carsickness group at baseline (Figure 3). Following training, symptom reduction in the MM-BCI group coincided with a normalization of this exponent, whereas the sham group showed no significant change in either measure (Figure 4 and Figure S2). These results confirm the occipito-parietal aperiodic exponent as a reliable neural signature for carsickness susceptibility and demonstrate that the MM-BCI training can effectively modulate it, suggesting an associated optimization of brain function. This finding advances prior EEG research on motion sickness, which has predominantly focused on band-limited oscillatory power [27], [28], [29]. Such approaches typically do not separate the aperiodic component, inherently conflate distinct neurophysiological processes [23]. This conflation, such as merging true oscillatory changes with broad-scale shifts in neural excitability, likely contributes to inconsistent findings and obscures specific neural mechanisms [30]. Supporting this view, our preliminary analysis showed that gamma relative power during car-riding state distinguished carsickness-susceptible individuals only when the aperiodic component was retained (Figure S3).

Aperiodic exponent is considered to reflect the brain’s responsivity to external stimuli, where a lower exponent indicates increased cortical excitability during the processing and integration of multimodal sensory inputs [31], [32]. Our finding of a significantly lower baseline occipito-parietal aperiodic exponent in susceptible individuals suggests a state of abnormally heightened excitability in key brain regions for integrating visual, vestibular, and proprioceptive inputs [33]. This neurophysiological trait may predispose them to experience carsickness symptoms when sensory conflicts arise during travel. The MM-BCI training ameliorates this by guiding users to redirect attentional resources away from internal discomfort and toward mindfulness meditation. This process effectively modulates the allocation of attention to sensory conflict. The subsequent normalization of the aperiodic exponent after repeated training, to a level indistinguishable from non-susceptible individuals, indicates that the intervention successfully down-regulates this aberrant excitability and reduces neural sensitivity to sensory conflict, thereby underpinning the treatment effect.

Several limitations of this study should be considered. First, although EEG is a powerful tool for assessing real-time brain activity, it provides limited information about functional connectivity between brain regions. Incorporating other neuroimaging techniques, such as functional magnetic resonance imaging (fMRI), could yield a more detailed, network-level understanding of the mechanisms underlying carsickness and its treatment. Second, the follow-up period in this study was limited to one month. Longer-term assessments are needed to evaluate the durability of the intervention’s effects and to determine whether periodic booster sessions are necessary to maintain symptom relief over time. Lastly, while we targeted carsickness in the present study, the potential treatment effect on other forms of motion sickness, such as seasickness or visually induced motion sickness, warrants further investigation.

In conclusion, this study demonstrates that MM-BCI training is a feasible and effective carsickness therapy in real-world settings. By enhancing mindfulness meditation with real-time EEG neurofeedback, the intervention produced significant treatment effect on carsickness and selectively modified occipito-parietal aperiodic activity associated with carsickness susceptibility. These findings advance our understanding of the neural mechanisms underlying carsickness and highlight the potential of lightweight, wearable BCI technology to increase resilience to motion-induced discomfort. More broadly, this research paves the way for using closed-loop neurofeedback to modulate multisensory brain dynamics, opening new avenues for improving cognitive control and promoting human well-being in everyday environments.

## 4 Methods

The core of this study was a randomized controlled trial designed to evaluate the treatment efficacy of the MM-BCI system on carsickness and to explore its underlying neural mechanisms. Within this framework, a cross-sectional comparison was embedded to provide a baseline reference for the neural signature associated with carsickness susceptibility. The study protocol was approved by South China Normal University Human Research Ethics Committee for Non-Clinical Faculties (Approval No. SCNU-BRR-2024-069) and was prospectively registered with the Chinese Clinical Trial Registry (Registration No. ChiCTR2500103519). All participants were free to withdraw from the study at will and provided written informed consent prior to enrolment.

### 4.1 Participants

Participants were recruited publicly via campus advertisements and online platforms. All applicants first completed a screening questionnaire comprising demographic information and the Motion Sickness Susceptibility Questionnaire shortened version (MSSQ-Short) [34]. Individuals who self-reported frequent discomfort or nausea when riding in cars and obtained an MSSQ-Short score greater than 11.3 (corresponding to a susceptibility percentile greater than 50%) were enrolled in the carsickness group. Those who self-reported never experiencing discomfort or nausea in cars and scored less than 4.1 on the MSSQ-Short (corresponding to a susceptibility percentile less than 20%) were enrolled in the non-carsickness group. Applicants were excluded if they had a history of vestibular nerve damage; traumatic brain injury; or diagnosed neurological, cardiovascular, musculoskeletal, or other significant active systemic diseases. Additional exclusion criteria included the presence of metal implants, a family history of psychiatric disorders, current use of psychotropic medications, or being pregnant or breastfeeding.

Following screening, a total of 60 eligible participants were included in the carsickness group and 31 in the non-carsickness group. Participants in the carsickness group were randomly allocated in a 1:1 ratio to either the MM-BCI group or the sham group using a computer-generated randomization sequence (Figure 1A). The allocation sequence was concealed and managed by a researcher not involved in participant recruitment or assessment. Participants in the carsickness group were blinded to their group assignment. Participants in the non-carsickness group did not undergo randomization and formed a separate baseline control cohort. The three groups did not differ significantly in age or gender distribution. As expected, both the MM-BCI and sham groups showed significantly higher MSSQ-Short scores compared to the non-carsickness group. Detailed participant characteristics are presented in Table 1.

**Table 1.** Demographic and motion sickness susceptibility characteristics of participants.

|  | MM-BCI group (N=30) | Sham group (N=30) | Non-carsickness group (N=31) | Statics | P |
| --- | --- | --- | --- | --- | --- |
| Age (year) | 21.86±0.40 | 21.80±0.31 | 21.96±0.48 | F(2,88)=0.043 | 0.957 |
| Gender | 11M/19F | 16M/14F | 19M/12F | X<sup>2</sup>(2)=3.837 | 0.147 |
| MSSQ-Short score | 31.90±1.47<sup>a</sup> | 32.56±1.66<sup>b</sup> | 2.19±0.24 | F(2,88)=186.794 | <0.001 |

<sup>a</sup> a significant difference compared to non-carsickness group (P<0.001, Benjamini-Hochberg correction)
<sup>b</sup> a significant difference compared to non-carsickness group (P<0.001, Benjamini-Hochberg correction)

### 4.2 Assessments

To assess carsickness severity, all participants were scheduled to complete three standardized assessments conducted respectively at pre-intervention (baseline), post-intervention, and one-month follow-up. Each assessment involved a one-hour real-world car ride experiment in Guangzhou, China, following a fixed route that included typical urban road elements, such as straight segments, turns, and traffic lights. A single vehicle and driver were employed throughout all assessments to minimize extraneous variability.

During the experiment, the experimenter occupied the front passenger seat, while the participant sat directly behind, in the rear passenger-side seat. The procedure began with a 3-minute resting state EEG recording prior to vehicle ignition, during which participants were instructed to remain quiet with their eyes open naturally. Following this, the car commenced the one-hour car-riding state. At the beginning and at 10-minute intervals thereafter, the experimenter prompted the participant to verbally report their current carsickness severity. Participants were instructed to remain alert and relaxed throughout the ride, performing no tasks other than providing these ratings. A printed version of MISC [24] was affixed to the back of the front passenger seat for participant reference. Participants matched their subjective experience to the symptom descriptors on the scale and reported the corresponding score.

In both the pre- and post-intervention assessments, EEG signals were synchronously recorded during the car-riding state using a 64-channel cap following the international 10–20 system, connected to a Synamps2 amplifier (Compumedics, Neuroscan, Inc., Australia) with a sampling rate of 1 kHz. During the experiment, electrode impedances were maintained below 5 kΩ, with the ground electrode placed on the forehead. Following the post-intervention assessment, participants in the MM-BCI and sham groups additionally completed a 7-point Likert scale item to evaluate the perceived effectiveness of the 10-week training for treating carsickness. Subsequently, a one-month follow-up assessment was conducted, which included only the subjective carsickness severity ratings. Four participants from each of the MM-BCI and sham groups were unable to participate in the follow-up due to scheduling conflicts, resulting in a final sample of N=26 per group for this assessment. Participants in the non-carsickness group underwent only the baseline assessment and did not participate in the subsequent intervention or follow-up.

### 4.3 Training sessions

Both the MM-BCI and sham groups completed a 10-week intervention, consisting of one session per week. Each weekly session included a one-hour real-world car-riding meditation training and two 30-minute at-home meditation training. During the car-riding meditation training, participants used their respective neurofeedback systems (the MM-BCI system delivering real-time neurofeedback vs. a sham feedback system) while performing mindfulness meditation. The audiovisual feedback interface was displayed on a screen fixed to the back of the front passenger seat. As in the pre- and post-intervention assessments, the experimenter prompted and recorded the participant’s verbally reported carsickness severity at the start and at 10-minute intervals throughout car riding. For the at-home meditation training, participants used the system to complete two 30-minute mindfulness meditation training outside the vehicle. The proposed MM-BCI system comprised three components: a wearable EEG headband, client software, and a graphical interface. The headband (HNNK, http://www.ihnnk.com/, Guangdong, China) primarily acquired EEG signals from the right prefrontal site (Fp2), with the ground and reference electrodes placed on the left and right temples, respectively. Dry electrodes were used, with impedance maintained below 5 kΩ. The EEG signals were transmitted wirelessly in real-time to the client software for decoding. The software hosted a pre-trained model that decoded the user’s EEG signals and quantified it into a meditation score ranging from 0 to 100 [13], [14], [35]. A higher score indicated a deeper, more focused meditative state. Furthermore, the software provided various audiovisual meditation scenes, each designed by a professional meditation instructor and accompanied by guided meditation instructions. Changes in these audiovisual scenes (e.g., enrichment of visual elements, increase in background sound volume) were dynamically modulated by the user’s real-time meditation score every second and fed back via the graphical interface.

The sham feedback system was identical in configuration to the MM-BCI system, with the key distinction that transitions in its audiovisual scenes were not controlled by the user’s real-time meditation score. Instead, the system employed a yoked-control design [36], whereby each participant from the sham group was randomly paired with a participant from the MM-BCI group. The sham group participant received an identical playback of the audiovisual feedback sequence that was generated by their paired MM-BCI participant’s EEG signals during a corresponding session, while still following the same guided instructions.

Prior to the commencement of the intervention in session 1, all participants in both the MM-BCI and sham groups received identical standardized training. This training introduced the benefits and core techniques of mindfulness meditation (e.g., maintaining focus on one’s breath) and instructed participants to actively adjust their meditative state in response to the audiovisual feedback provided by their respective systems.

### 4.4 Data analysis

#### 4.4.1 Behavioral data

Following the post-intervention assessment, participants in the MM-BCI and sham groups rated the statement, “The 10-week training was an effective therapy on carsickness”, on a 7-point Likert scale ranging from 1 (strongly agree) to 7 (strongly disagree). For descriptive analysis, responses were categorized, with scores of 1 to 3 (strongly agree, agree, somewhat agree) classified as a positive evaluation of the training’s efficacy. The percentage of participants providing a positive evaluation was calculated for each group.

To quantify carsickness severity during the car-riding state, participants verbally reported their symptom levels using MISC at seven time points, including immediately upon departure (0 min) and at 10-minute intervals thereafter. For each assessment, a mean carsickness severity score was calculated by averaging the six MISC scores obtained from the 10- to 60-minute intervals. To measure the treatment effect, the MISC relative change (Δ*MISC*) was computed. This metric was defined, for each participant, as the difference between the post-intervention and pre-intervention mean MISC scores, divided by the pre-intervention mean MISC score, as shown in Equation (1).

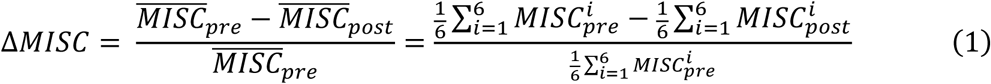

#### 4.4.2 EEG data

##### (1) Pre-processing

For pre- and post-intervention assessments, the multi-channels EEG data were first pre-processed by the following steps. (1) The EEG data were resampled to 250 Hz. (2) The 50 Hz a.c. line noise artifact was removed by a notch filter. (3) Slow drifts in the EEG recordings were removed using a 1 Hz high-pass filter. (4) The artifacts with high-frequency components were removed using a 100 Hz low-pass filter. (5) Bad channels were rejected based on local outlier factor algorithm [37]. The rejected bad channels were then interpolated from the EEG of adjacent channels via the spherical spline interpolation [38]. (6) Remaining artefacts were removed using independent component analysis [39]. Independent components related to the muscle artifact, ocular artifact and electrocardiographic artifacts were automatically rejected using a pattern classifier trained on expert-labelled independent components from another independent EEG dataset [40]. (7) EEG data were re-referenced to the common average. Following preprocessing, EEG data were segmented relative to the experimental timeline. For each assessment, the following were extracted: (1) a single 3-minute segment of resting state EEG data, and (2) from the car-riding state, six 3-minute segments, each corresponding to the period immediately preceding a verbal symptom report (i.e., spanning the 10- to 60-minute intervals).

##### (2) Spectral Parameterization for Aperiodic Component

The aperiodic component of the EEG data was parameterized using a validated spectral decomposition approach [23]. The power spectral density (PSD) was first estimated using the Welch method, with a Hanning window applied to 2-second segments and 50% overlap, yielding a frequency resolution of 0.5 Hz. Subsequent analysis was restricted to the frequency range of 1 to 45 Hz.

To decompose the PSD into periodic and aperiodic components, we employed the “Fitting Oscillations & One Over F” (FOOOF) algorithm [23]. This algorithm models the PSD in log-power space as a combination of an aperiodic component and a sum of oscillatory peaks modeled as Gaussian functions. The model is formally defined by the equation:

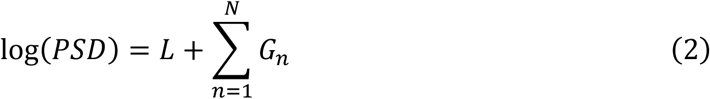

where *L* represents the aperiodic component and 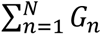 represents the sum of *N* Gaussian functions fitting the periodic oscillations. The aperiodic component *L* was fit in “fixed” mode, corresponding to a linear fit in log-log space, and is described by:

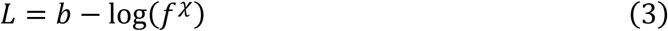

where *b* is the offset and *χ* is the exponent, which characterizes the slope of the aperiodic component. A steeper slope (higher *χ*) indicates a faster decay of power with increasing frequency. The parameterization settings were configured as follows to optimize the fit for our data: peak width limits: [1.0, 8.0], max number of peaks: 6; minimum peak height: 0.1; peak threshold: 2.

The primary output of interest for subsequent analysis was the aperiodic exponent (*χ*), which quantifies the slope of the 1/f-like background activity. A single mean aperiodic exponent was calculated for each state. The resting state value was derived directly from its single segment. For the car-riding state, the six segment values were averaged to obtain one mean per assessment. Finally, the intervention-related neural change for each state was quantified as the aperiodic exponent relative change, calculated by the same formula applied to the behavioral data (Equation (1)). For the correlation analysis between the aperiodic exponent relative change and the MISC relative change, outliers were identified and excluded based on a threshold of 2.5 standard deviations from the group mean for both variables. This outlier removal procedure led to the exclusion of 2 participants from the MM-BCI group and 1 participant from the sham group, ensuring the robustness of the correlation results.

##### (3) Headband EEG data processing

For car-riding meditation training, the single-channel headband EEG data were first pre-processed by notch filtering at 50 Hz and high-pass filtering at 1 Hz. Data segmentation was performed identically to the multi-channel EEG data. Specifically, in each session, six 3-minute car-riding state segments (each preceding a symptom report) were extracted. The power spectral density for each segment was estimated using the Welch method. The absolute spectral power in the theta (4-7 Hz) and alpha (8-13 Hz) frequency bands was then computed. The theta/alpha power ratio was then calculated for each segment as the ratio of theta to alpha power. This feature was selected based on previous studies demonstrating its relevance to attentional states, where a lower frontal theta/alpha ratio is associated with a meditative state compared to mind-wandering, and the ratio decreases with increasing meditation depth [25], [26]. Finally, the six theta/alpha ratio values from the car-riding meditation training were averaged for each participant, yielding a single mean theta/alpha ratio feature for the car-riding meditation training per session.

#### 4.4.3 Statistical analysis

Data distributions are reported as mean ± s.e.m. All statistical tests were two-tailed. The threshold for statistical significance was set at P < 0.05. For each significant effect, the analyses of variance (ANOVA) and t test effect sizes were measured via ηp² and Cohen’d respectively. For analyses involving multiple comparisons, p-values were adjusted using the Benjamini-Hochberg correction.

To examine the effects of the intervention on carsickness severity, MISC scores were analyzed using two-way repeated-measures ANOVAs. A 2 (group: MM-BCI, sham) × 3 (time: pre-intervention, post-intervention, 1-month follow-up) ANOVA was conducted to assess the main and interaction effects across the primary assessment time points. Separately, to analyze the progression during the car-riding meditation training, MISC scores from the weekly car-riding meditation training were submitted to a 2 (group) × 10 (Week) ANOVA.

For EEG-derived features (e.g., aperiodic exponent), between-group comparisons (MM-BCI vs. sham vs. non-carsickness) at pre- and post-intervention were assessed separately using one-way ANOVAs. Within-group changes from pre- to post-intervention were evaluated using paired-samples t-tests for the MM-BCI and sham groups independently.

To identify brain regions associated with carsickness susceptibility, a cluster-based permutation test [41], [42] was applied to the baseline (pre-intervention) aperiodic exponent across all electrodes. The statistical model for the cluster-forming stage was a one-way ANOVA, which assessed differences across all three groups (MM-BCI, sham and non-carsickness) simultaneously. This non-parametric method controls the family-wise error rate in the context of multiple spatial comparisons. The analysis was implemented with the number of permutations set to 5000 and the cluster-forming threshold corresponding to P < 0.01. Features from electrodes within significant clusters were averaged to create a composite EEG signature for subsequent analyses. The relationship between neural change and symptom relief was examined by calculating the Pearson correlation coefficient between the aperiodic exponent relative change and the MISC relative change.

The cluster-based permutation analysis was implemented using the MNE-Python library (version 1.10). All other statistical analyses were performed using IBM SPSS Statistics (Version 27.0).

## Data Availability

The data that support the findings of this study are available from the corresponding authors upon reasonable request.

## 5 Acknowledgements

We thank all participants of the study. This work was supported in part by Brain Science and Brain-like Intelligence Technology-National Science and Technology Major Project under Grant 2022ZD0208900; in part by the Key Research and Development Program of Guangdong Province, China under Grant 2018B030339001; in part by the Guangzhou Talent Plan under Grant 2024D02J0008; in part by National Key Research and Development Program of China under 2025YFE0213500; in part by Guangdong Natural Science Foundation under Grant 2024A1515011690; in part by the Guangdong Talent Plan under Grant 2023QN100110.

## 6 Author contribution

Y.L. was fully responsible for the conceptualization and supervision of this study. Y.L. and

J.Z. designed and organized the experiments. J.Z. and Y.C. collected the data. J.Z., Z.W., Q.H. and Y.L. jointly analyzed the data. J.Z., Z.W., Q.H. and Y.L. jointly wrote the paper.

## 7 Competing interests

The authors declare that they have no competing interests.

## Notes

### Competing Interest Statement

The authors have declared no competing interest.

### Clinical Trial

ChiCTR2500103519

### Author Declarations

The study protocol was approved by South China Normal University Human Research Ethics Committee for Non-Clinical Faculties (Approval No. SCNU-BRR-2024-069).

