## Supplementary Materials for "Carsickness Therapy Based on Brain–Computer Interface Enhanced Mindfulness Meditation Training"

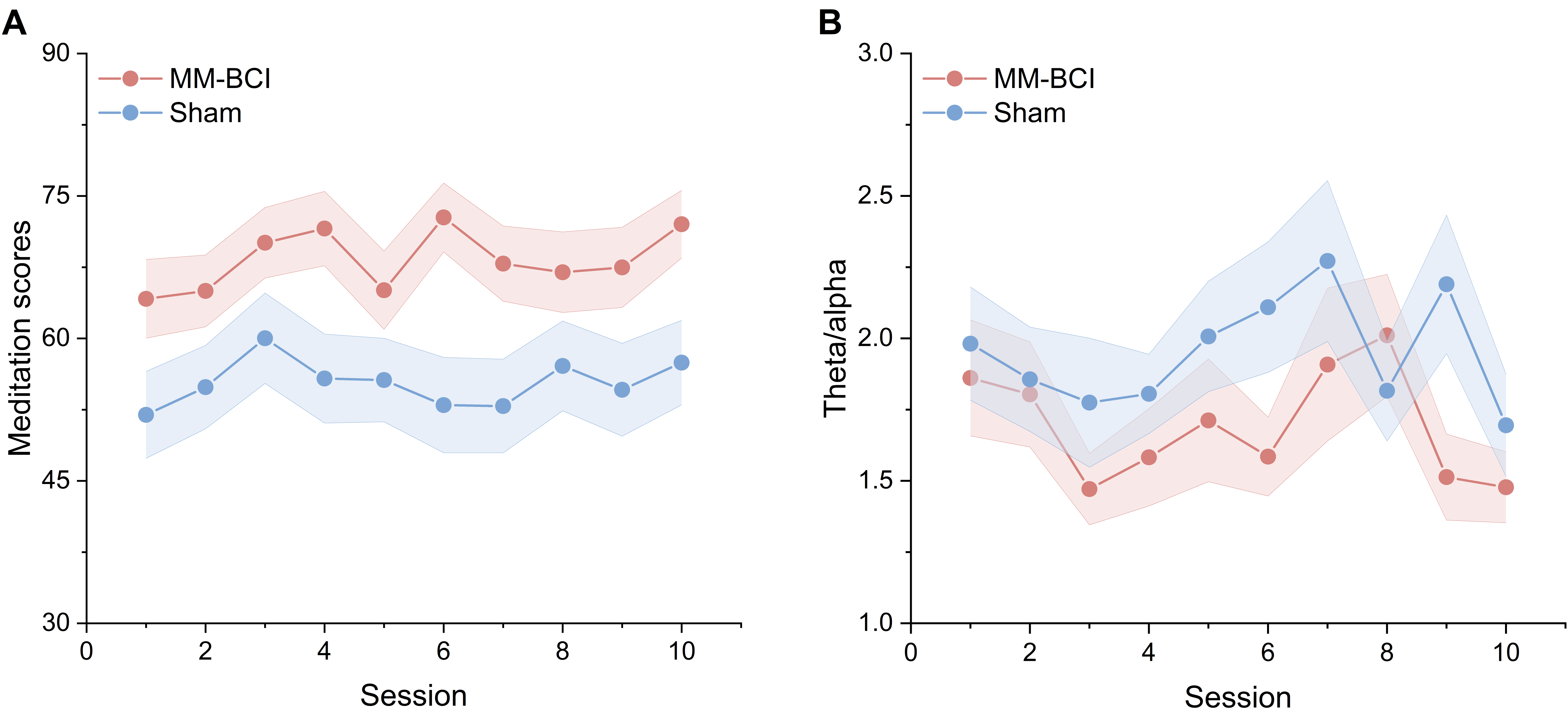

Figure S1. The headband EEG results during car-riding meditation training. The averaged meditation scores (A) and theta/alpha ratio (B) for all participants within each group from session 1 to 10. The shadow represents the standard error of the mean (s.e.m).

The MM-BCI group demonstrated a superior ability to maintain focused attention meditation, as evidenced by both meditation scores and neural metrics. Analysis of the real-time meditation scores (Figure S1A) yielded a significant main effect of group (F(1, 580) = 44.944, P < 0.001, ηp² = 0.072), with the MM-BCI group sustaining higher scores throughout. The main effect of session (F(9, 580) = 0.546, P = 0.841) and the group × session interaction (F(9, 580) = 0.296, P = 0.976) were not significant.

This finding was further corroborated by the physiological theta/alpha power ratio derived from the single-channel headband EEG (Figure S1B). Prior to analysis, samples exceeding ±2.5 standard deviations from the weekly group mean were excluded as outliers, resulting in the exclusion of 19 samples across all groups and sessions. A two-way ANOVA on this metric also revealed a significant main effect of group (F(1, 561) = 8.532, P = 0.004, ηp² = 0.015), with the MM-BCI group maintaining a lower ratio, indicative of a more focused meditative state. Neither the main effect of session (F(9, 561) = 1.170, P = 0.311) nor the interaction (F(9, 561) = 0.750, P = 0.663) reached significance.

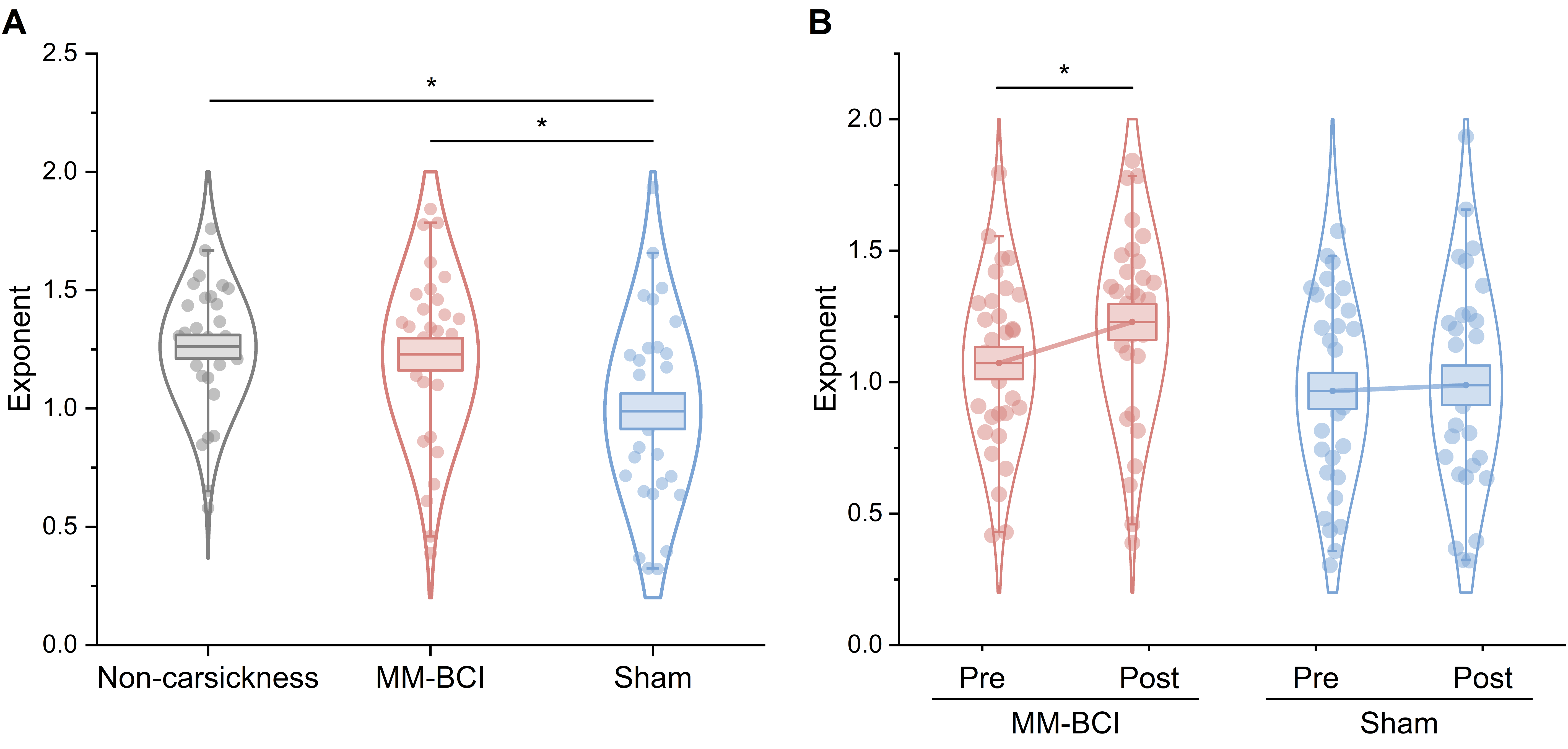

Figure S2. Post-intervention results of the occipito-parietal aperiodic exponent during the resting state. A, Mean aperiodic exponent (averaged across significant electrode cluster O1, Oz, PO5) for the MM-BCI and sham groups at post-intervention assessment against the baseline of non-carsickness group. B, Mean aperiodic exponent for the MM-BCI and sham groups at pre- and post-intervention assessment. The center line represents the mean, the box indicates s.e.m, and the whiskers extend to the 5th and 95th percentiles. *P < 0.05.

During the resting state, a one-way ANOVA on post-intervention aperiodic exponent indicated a significant group effect (F(2, 88) = 5.255, P = 0.007, ηp² = 0.107; Figure S2A). Post-hoc independent-samples t-tests with Benjamini-Hochberg correction showed the aperiodic exponent of the MM-BCI group was significantly higher than the sham group (t(58) = 2.369, P = 0.032, Cohen’d = 0.612) and comparable to the non-carsickness baseline (t(59) = 0.382, P = 0.704). The sham group remained significantly reduced relative to the non-carsickness group (t(59) = -3.049, P = 0.010, Cohen’d = -0.781). Paired-samples t-tests examining within-group changes from pre- to post-intervention revealed a significant improvement in aperiodic exponent for the MM-BCI group (t(29) = 2.117, P = 0.043, Cohen’s d = 0.386; Figure S2B), whereas the sham group showed no significant change (t(29) = 0.346, P = 0.732).

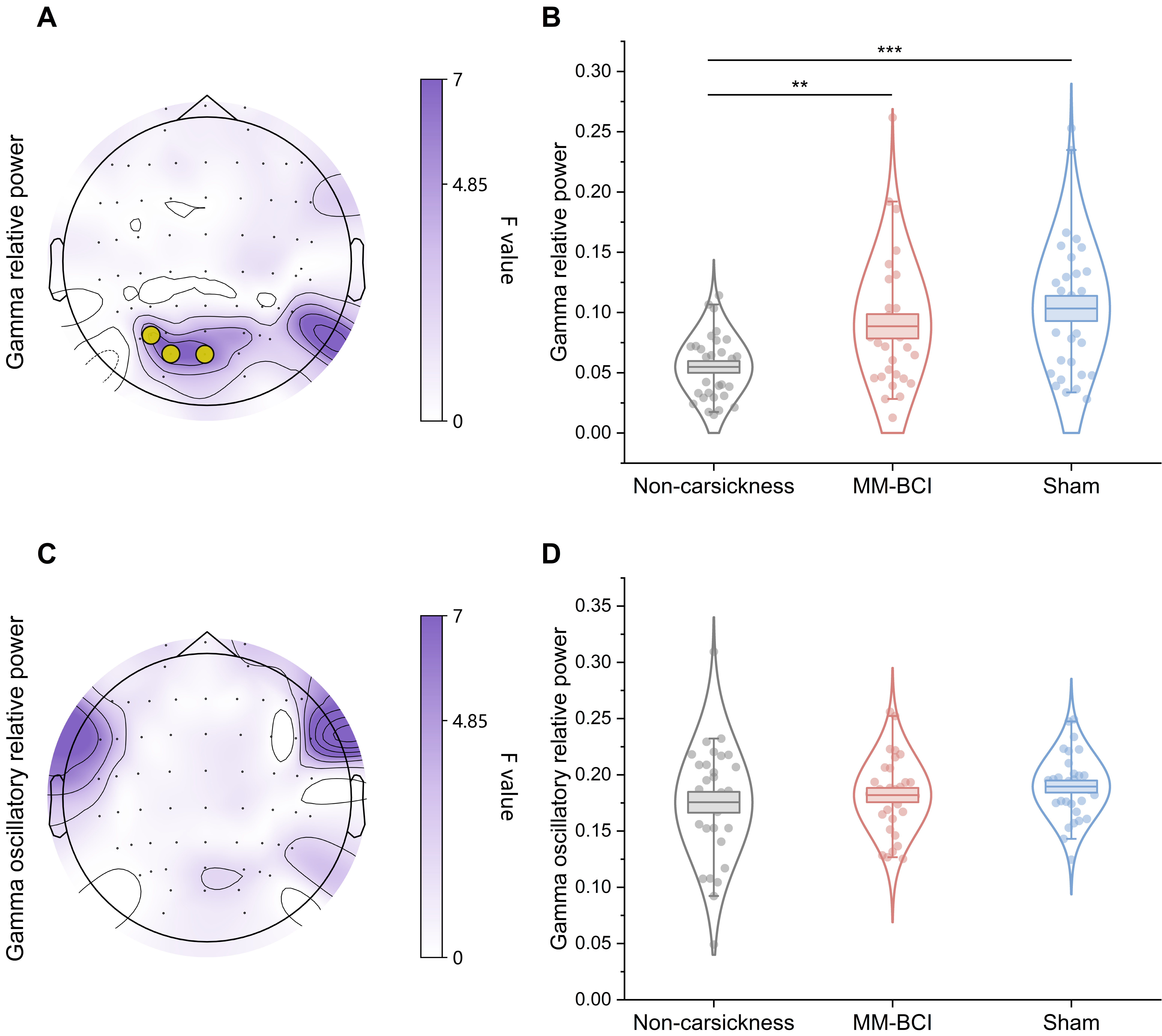

Figure S3. Baseline (pre-intervention) differences in gamma relative power associated with carsickness susceptibility during the car-riding state. A, Topographic map of the F-statistic from a one-way ANOVA comparing the aperiodic exponent across the non-carsickness, MM-BCI, and sham groups during the car-riding state. Electrodes forming a significant cluster (O1, Oz, PO5) after cluster-based permutation correction (P_cluster_ = 0.029, ηp^2^ = 0.155) are outlined in yellow circle. B, The mean aperiodic exponent averaged across the significant electrode cluster identified in (A) for individuals in each group during the car-riding state. C, D, Corresponding topographic map of the F-statistic (C) and group-level boxplots (D) for the gamma oscillatory relative power after removing aperiodic component. The center line represents the mean, the box indicates s.e.m, and the whiskers extend to the 5th and 95th percentiles. **P < 0.01, ***P < 0.001.

The cluster-based permutation test (one-way ANOVA model), with a threshold set at P = 0.01(equivalent to F = 4.85), applied to the gamma relative power during the car-riding state revealed one significant cluster comprising 3 electrodes (O1, Oz, PO5; F_cluster_(2, 88) = 6.785, P_cluster_= 0.029, ηp^2^ = 0.155; Figure S3A). Post-hoc independent-samples t-tests with Benjamini-Hochberg correction were conducted to compare the averaged power from this cluster between groups. The analysis showed no significant difference between the MM-BCI and sham groups (t(58) = 1.015, P = 0.314; Figure S3B). In contrast, both the MM-BCI group (t(59) = 3.033, P = 0.005, Cohen’d = -0.777) and the sham group (t(59) = 4.243, P < 0.001, Cohen’d = 1.086) exhibited significantly higher gamma relative power in this occipito-parietal cluster compared to the non-carsickness group.

We conducted an identical analysis on gamma oscillatory relative power, which was derived by removing the aperiodic component. However, no significant cluster emerged (Figure S3C), and the average power across electrodes O1, Oz, and PO5 revealed no significant differences among the three groups (F(2, 88) = 0.932, P = 0.398; Figure S3D).

Furthermore, identical analyses performed on the relative power of other bands (including delta, theta, alpha and beta) revealed no significant differences between the non-carsickness and carsickness groups.
